# Synergistic effects of cardiovascular health and social isolation on adverse pregnancy outcomes

**DOI:** 10.1101/2024.07.05.24309978

**Authors:** Hisashi Ohseto, Mami Ishikuro, Geng Chen, Ippei Takahashi, Genki Shinoda, Aoi Noda, Keiko Murakami, Masatsugu Orui, Noriyuki Iwama, Masahiro Kikuya, Hirohito Metoki, Atsushi Hozawa, Taku Obara, Shinichi Kuriyama

**Affiliations:** Graduate School of Medicine, Tohoku University, Sendai, Miyagi, Japan; Tohoku Medical Megabank Organization, Tohoku University, Sendai, Miyagi, Japan; Tohoku University Hospital, Tohoku University, Sendai, Miyagi, Japan; Graduate School of Medicine, Teikyo University, Itabashi-ku, Tokyo, Japan; Graduate School of Medicine, Tohoku Medical and Pharmaceutical University, Sendai, Miyagi, Japan; International Research Institute of Disaster Science, Tohoku University, Sendai, Miyagi, Japan

**Keywords:** Cohort study, Gestational diabetes, Health disparities, Income, Life’s Essential 8, Preeclampsia, Prenatal care, Preterm birth, Psychological distress, Small for gestational age

## Abstract

**Background:** Adverse pregnancy outcomes affect approximately 20% pregnant women, and their incidence is increasing.

**Objective:** To investigate the effect of cardiovascular health during pregnancy on adverse pregnancy outcomes and the effect modification by psychological distress, social isolation, and income.

**Study Design:** We analyzed data from 14,930 pregnant women in the Tohoku Medical Megabank Project Birth and Three-Generation Cohort Study. Cardiovascular health status during pregnancy was assessed using the eight components of Life’s Essential 8 as proposed by the American Heart Association, including diet, physical activity, nicotine exposure, sleep health, body mass index, blood lipids, blood glucose, and blood pressure. Adverse pregnancy outcomes were defined as composite outcomes of preeclampsia, gestational diabetes mellitus, preterm birth, and small for gestational age. Using logistic regression analyses, we examined the associations between cardiovascular health and adverse pregnancy outcomes, preeclampsia, gestational diabetes mellitus, preterm birth, small for gestational age, large for gestational age, low birth weight, and neonatal intensive care unit admission. Interactions with psychological distress, social isolation, and income were examined.

**Results:** The numbers of participants with high, moderate, and low cardiovascular health status were 2,891 (19.4%), 11,498 (77.0%), and 541 (3.6%), respectively. Moderate and low cardiovascular health status were positively associated with adverse pregnancy outcomes (odds ratio and 95% confidence interval: 1.17 (10.04 to 1.32) and 2.64 (2.13 to 3.27), respectively). Low cardiovascular health status was also associated with a higher prevalence of preeclampsia, gestational diabetes mellitus, preterm birth, large for gestational age, and neonatal intensive care unit admission, and lower prevalence of small for gestational age. Among pregnant women with low cardiovascular health status, those who reported social isolation had a higher prevalence of adverse pregnancy outcomes than did those without social isolation (36.4% vs. 27.4%). However, this difference was attenuated for pregnant women with high cardiovascular health status (13.6% vs. 13.1%).

**Conclusions:** Cardiovascular health status assessed using Life’s Essential 8 may be useful for assessing the risk of adverse pregnancy outcomes. Socially isolated pregnant women are more vulnerable to the effects of low cardiovascular health status; thus, they should be prioritized for access to primary care, lifestyle education, and appropriate pharmacotherapy.

**Tweetable Statement:** Cardiovascular health impacts pregnancy outcomes. Social isolation exacerbates these risks. Improving cardiovascular health, particularly in socially isolated women, is crucial to reducing health disparities.

**AJOG at a Glance:** *Why was this study conducted?:*

- Adverse pregnancy outcomes affect approximately 20% pregnant women and are increasing. We hypothesized that preventive strategies employed for cardiovascular diseases can be applied to adverse pregnancy outcomes.

*What are the key findings?:*

- Pregnant women with low cardiovascular health status had an increased risk of adverse pregnancy outcomes, preeclampsia, gestational diabetes mellitus, preterm birth, large for gestational age, and neonatal intensive care unit admission.
- In addition, socially isolated pregnant women were more affected by low cardiovascular health status and faced a greater risk of adverse pregnancy outcomes.

*What does this study add to what is already known?:*

- Improving cardiovascular health during pregnancy, especially for socially isolated women, is necessary to minimize health disparities.

## Introduction

Adverse pregnancy outcomes (APOs), which encompass unfavorable events or complications occurring during pregnancy, delivery, or postpartum, affect approximately 20% pregnant women and are increasing.^1, 2^ Following delivery, APOs can progress to cardiovascular disease (CVD)^3–5^ and mortality.^6^ Placental formation and cardiometabolic factors play crucial roles in APO development,^7, 8^ and their pathogenic similarity to CVDs has led to the characterization of pregnancy as a “stress test for CVD.”^9^ Risk factors for APOs, such as obesity,^10^ poor sleep quality,^11^ and poor dietary habits^12^ are also recognized as risk factors for CVD.^13^ Therefore, it is expected that similar preventive strategies employed for CVD can be applied to APOs.

In 2022, Life’s Essential 8 (LE8) proposed by the American Heart Association,^13^ which is an approach grounded in cardiovascular health (CVH) for disease prevention and health promotion. This concept emphasizes individual health while addressing existing CVD and associated risk factors. LE8 serves as an operational metric for CVH that includes eight components: diet, physical activity (PA), nicotine exposure, sleep health, body mass index (BMI), blood lipids, blood glucose, and blood pressure (BP). In contrast to Life’s Simple 7 (LS7), a prior version of CVH metrics proposed in 2010,^13^ sleep health was introduced as a new component in 2022, with all eight CVH components being rescaled to continuous variables. To date, over 2,500 studies have referenced original articles describing LS7,^13^ and evidence on LE8 is growing.^14, 15^ A meta-analysis revealed associations between CVH, and CVD incidence, CVD mortality, and all-cause mortality.^16^ Furthermore, these associations have been observed in adolescents and young adults.^17^ Therefore, we hypothesized that LE8 may be relevant in a wide range of APOs due to shared characteristics between CVD and APOs.

The integration of LE8 into antenatal care could enhance maternal health. Many previous studies solely quantified the impact of a single component of CVH, neglecting its complex interconnections. LE8 provides a comprehensive overview of maternal CVH during pregnancy and is useful for assessment in clinical scenarios where a single factor (e.g., smoking cessation) is related to others (e.g., weight gain or poor dietary habits).^18^ To date, no study has investigated the relationship between LE8 during pregnancy and APOs. However, one study^19^ exploring the link between LS7, earlier CVH metrics, and APOs demonstrated that improved CVH was associated with better pregnancy outcomes. Notably, this study utilized only five of seven components, and it remains unclear whether a comprehensive CVH assessment using LE8 is beneficial in antenatal care. Furthermore, the clinical significance of psychological health and the social determinants involved in effect modification cannot be overlooked. They have a solid clinical basis and underpin all the CVH metrics, interacting bidirectionally.^13, 20, 21^ Previous research has revealed that poor CVH was related to depressive symptoms^22, 23^ and social isolation.^24^ Another study found an effect modification by CVH on the relationship between socioeconomic status and life expectancy.^20^ To date, no study has investigated effect modification by psychological health and social determinants on the relationship between CVH and APOs.

In this study, we aim to investigate the effect of CVH assessed using LE8 on APOs in 14,930 pregnant Japanese women. We will also explore the effect modification by psychological distress, social isolation, and income.

## Materials and Methods

### Participants

Between 2013 and 2017, the Tohoku Medical Megabank Project Birth and Three-Generation (TMM BirThree) Cohort Study^25, 26^ recruited 23,406 pregnant women from more than 50 obstetric clinics and hospitals in Miyagi Prefecture, Japan. Participants who withdrew consent (n=232), those with multiple births (n=313), those who delivered before or at 32 weeks of gestation (n=223), those who had their blood samples drawn after 32 weeks of gestation (n=3,724), and those with missing data on delivery status (n=965) were excluded. A cutoff of 32 weeks of gestation was chosen to capture cardiovascular changes on transition from the second to the third trimester and to prevent pregnancy complications from affecting the CVH status. Additionally, this cutoff value aligned with those in previous studies.^19, 27^ Participants that were involved in the TMM BirThree Cohort Study multiple number of times were identified, and only their first valid data were used in this study (n=620). Finally, participants with missing CVH metrics and covariates were excluded (n=3,391), leaving 14,930 participants for the main analysis. The study flowchart is detailed in Supplementary Figure 1. Ethical approval was obtained from the Ethics Committee of the Tohoku Medical Megabank Organization (2013-1-103-1), and all participants provided written informed consent for study participation.

### CVH definitions

Dietary quality was assessed using the 8-item Japanese Diet Index, which assesses adherence to the Japanese diet and is related to CVD mortality in the Japanese population.^28, 29^ Food intake was estimated using a food frequency questionnaire.^30^ PA, nicotine exposure, and sleep health were assessed during pregnancy using self-reported questionnaires. Pre-pregnancy BMI and BP before 20 weeks of gestation were obtained from medical records during antenatal care. Blood lipid and glucose levels before or at 32 weeks of gestation were determined from blood samples collected in the TMM BirThree Cohort Study.^25^ Each LE8 component was rated on a scale from 0 (least healthy) to 100 (most healthy), with detailed information provided in Supplementary Table 1. The overall CVH score was the unweighted average of all eight components.^13^ Overall and component CVH scores were used to categorize participants into three levels: high (80–100), moderate (50–79), and low (0–49), as outlined in the original article.^13^

### Outcomes

APOs were defined as composite outcomes comprising preeclampsia (PE), gestational diabetes mellitus (GDM), preterm birth (PTB), and small for gestational age (SGA), based on their pathophysiological relevance to CVH and their clinical impact.^5, 31^ A diagnosis of PE was established based on the American College of Obstetricians and Gynecologists guidelines,^32^ utilizing medical records of antenatal care. GDM was identified from the medical records at delivery. To ascertain PTB and SGA, neonatal gestational age at delivery and birth weight were obtained from medical records at delivery. Additional neonatal outcomes, such as low birth weight (LBW), large for gestational age (LGA), and neonatal intensive care unit (NICU) admission, were identified from the medical records at delivery.

### Covariates

Covariates included maternal age at conception (≥35 years old or not), alcohol consumption during pregnancy, conception via in vitro fertilization (IVF), parity (primipara or not), psychological distress during pregnancy, social isolation during pregnancy, and household income (≤4 million yen or not). Age at conception and parity were obtained from medical records during antenatal care, while the remaining variables were self-reported during pregnancy. Psychological distress was evaluated using the Kessler Psychological Distress Scale (K6),^33^ comprising six items rated on a 5-point Likert scale. The total score ranged from 0 to 24, with a score of ≥9 indicating psychological distress. Social isolation was evaluated using the Lubben Social Network Scale (LSNS-6),^34^ which consists of six items rated on a 6-point Likert scale. The total score ranged from 0 to 30, with a score of ≤11 indicating social isolation. In addition, the LSNS-6 has two subscales: family and friends. Each subscale score ranged from 0 to 15, and a score of ≤5 was considered social isolation (Supplementary Table 2).^34^

### Statistical analysis

Baseline characteristics, CVH scores, and study outcomes were compared across the overall CVH levels and according to the APO status. Continuous variables were analyzed using analysis of variance or student’s t-test, while categorical variables were analyzed using the chi-square test. Pearson’s correlation coefficients were calculated for all combinations of component CVH scores.

The association between overall and component CVH levels and study outcomes was explored using multiple logistic regression analysis, employing high CVH levels as references and adjusting for covariates. Interactions between overall CVH levels and psychological distress, social isolation, and income levels were examined. Given that a significant interaction was observed with social isolation, further subgroup analyses were conducted for the subscales and each item of the LSNS-6 (Supplementary Table 2).

Sensitivity analyses were performed to assess the robustness of the results. First, due to some missing values not satisfying the “missing completely at random” assumption,^35^ we conducted association analyses after imputing missing values in CVH metrics and covariates using multiple imputation by chain equations with five iterations^36^ in a population of 18,321 eligible participants. Second, to rule out the potential of reverse causality between CVH and APOs, association analyses were conducted solely on participants whose blood samples were collected before or at 20 weeks of gestation (Supplementary Figure 2). Third, we excluded participants with pre-pregnancy chronic hypertension (CH) (n=579) or pre-pregnancy diabetes mellitus (DM) (n=71) from the main analysis. This exclusion aimed to prevent an overestimation of the effect of overall CVH on APOs, given the high risk of PE for patients with pre-pregnancy CH and the consequent diagnosis of GDM for those with pre-pregnancy DM.

A threshold p-value of <0.05 was adopted for detecting statistical significance for the association analyses, while a significance level of P < 0.2 was applied for the interaction term. All analyses were conducted using the R software version 4.1.2.

## Results

Among the 14,930 study participants, 2,891 (19.4%), 11,498 (77.0%), and 541 (3.6%) had high, moderate, and low overall CVH levels, respectively (Table 1). Among participants with high, moderate, and low overall CVH levels, 380 (13.1%), 1,772 (15.4%), and 162 (29.9%), respectively, had APOs (Supplementary Table 3). Pregnant women with APOs had lower overall CVH scores as well as lower scores for nicotine exposure, sleep health, BMI, blood glucose, and BP (Supplementary Table 4). As shown in Supplementary Figure 3, CVH scores for individual components were observed to correlate positively with each other. However, significant negative correlations existed between PA and nicotine exposure, PA and sleep health, PA and BMI, and diet and blood glucose CVH scores.

**Table 1.** Baseline characteristics of the complete cases in the TMM BirThree Cohort Study by overall CVH levels.

|  | High overall<br>CVH<br>n = 2,891 | Moderate<br>overall CVH<br>n = 11,498 | Low overall<br>CVH<br>n = 541 | P-value |
| --- | --- | --- | --- | --- |
| Advanced maternal age at<br>conception, % | 655 (22.7) | 3,092 (26.9) | 177 (32.7) | <0.001 |
| Low income, % | 875 (30.3) | 4,237 (36.8) | 269 (49.7) | <0.001 |
| Alcohol consumption during<br>pregnancy, % | 590 (20.4) | 2,303 (20.0) | 81 (15.0) | 0.012 |
| Psychological distress, % | 225 (7.8) | 1,237 (10.8) | 100 (18.5) | <0.001 |
| Social isolation, % | 470 (16.3) | 2,286 (19.9) | 154 (28.5) | <0.001 |
| Conception via IVF, % | 110 (3.8) | 611 (5.3) | 33 (6.1) | 0.002 |
| Primipara, % | 964 (33.3) | 4,698 (40.9) | 224 (41.4) | <0.001 |
| GA at blood sampling, day | 125.8 ± 30.2 | 138.3 ± 33.7 | 147.8 ± 34.6 | <0.001 |
| GA at BP measurement, day | 79.2 ± 14.8 | 79.4 ± 15.4 | 81.4 ± 14.8 | 0.006 |
| **CVH scores** |  |  |  |  |
| Overall CVH score | 85.0 ± 4.4 | 67.7 ± 7.3 | 44.4 ± 5.0 | <0.001 |
| Diet score | 69.9 ± 26.3 | 44.2 ± 32.5 | 22.7 ± 27.5 | <0.001 |
| PA score | 59.0 ± 45.3 | 16.7 ± 34.7 | 6.0 ± 21.9 | <0.001 |
| Nicotine exposure score | 88.3 ± 21.0 | 69.3 ± 33.8 | 33.2 ± 34.2 | <0.001 |
| Sleep health score | 83.5 ± 21.2 | 71.2 ± 26.6 | 48.5 ± 29.9 | <0.001 |
| BMI score | 96.9 ± 11.3 | 85.8 ± 26.2 | 42.4 ± 32.6 | <0.001 |
| Blood lipids score | 86.0 ± 22.5 | 65.9 ± 29.3 | 42.2 ± 26.2 | <0.001 |
| Blood glucose score | 99.8 ± 2.5 | 99.3 ± 5.9 | 93.6 ± 17.4 | <0.001 |
| BP score | 96.5 ± 10.5 | 89.3 ± 18.6 | 66.9 ± 26.6 | <0.001 |

CVH, cardiovascular health; IVF, in vitro fertilization; GA, gestational age; BP, blood
pressure; PA, physical activity; BMI, body mass index.
The overall CVH score was used to categorize participants into three groups: high (80–100), moderate (50–79), and low (0–49).
Data are shown as mean ± standard deviation for continuous variables and n (%) for categorical variables.
P-values were calculated using analysis of variance for continuous variables and the chi- square test for categorical variables.

In logistic regression analyses with adjustment (Table 2), moderate and low CVH levels were associated with APOs with odds ratios (OR) of 1.17 (95% CI: 1.04–1.32) and 2.64 (95% CI: 2.13 to 3.27), respectively (P for trend < 0.001). Low CVH levels were also associated with a higher prevalence of PE, GDM, PTB, LGA, and NICU admission and a lower prevalence of SGA. Using a heatmap, Figure 1 shows the results of logistic regression analyses with adjustments between eight component CVH levels, APOs, and other study outcomes. Low nicotine exposure, BMI, blood glucose, and BP CVH levels were positively associated with APOs. Low sleep health CVH levels, which was newly considered in LE8, were positively associated with GDM, SGA, and LBW, while diet and PA CVH were largely unrelated to the study outcomes. Low BMI, blood lipids, blood glucose, and BP CVH levels tended to be positively associated with the study outcomes, except that low BMI and blood lipids CVH levels were negatively associated with SGA and LBW.

**Figure 1.**
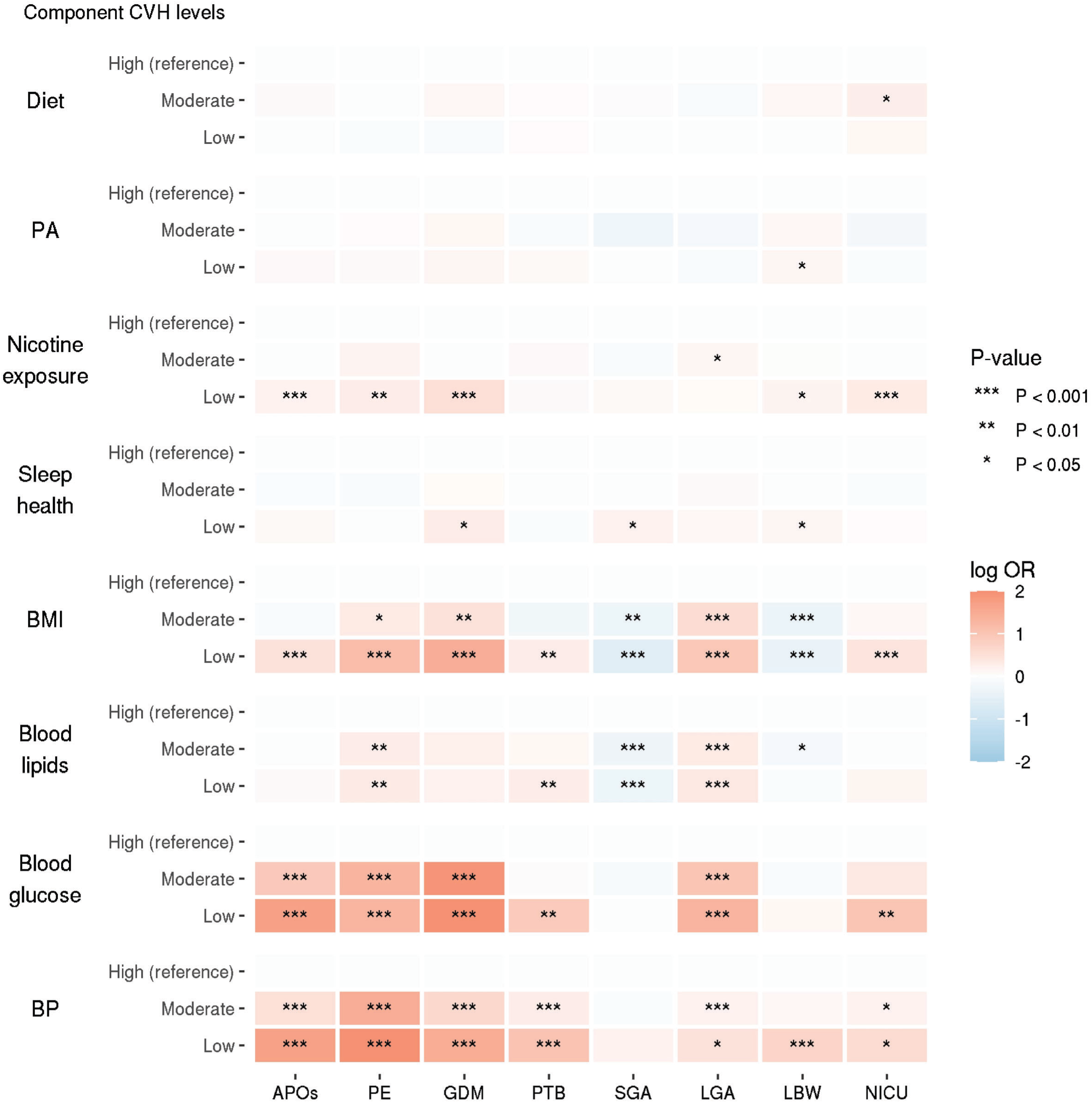
Heatmap showing associations between component CVH levels and study outcomes. The heatmap represents Log transformed ORs from multiple logistic regression analyses between component CVH levels and study outcomes. Adjustments were made for maternal age at conception, alcohol consumption during pregnancy, conception via in vitro fertilization, parity, psychological distress during pregnancy, social isolation during pregnancy, and household income. High CVH levels were references. Red, blue, and grey indicate positive, negative, and no association, respectively. Darker colors indicate stronger associations. The asterisk indicates the p-value. For clarity, extreme Log OR values are presented as 2 or -2 when their absolute values are 2 or more. CVH, cardiovascular health; OR, odds ratio; PA, physical activity; BMI, body mass index; BP, blood pressure; APOs, adverse pregnancy outcomes; PE, preeclampsia; GDM, gestational diabetes mellitus; PTB, preterm birth; SGA, small for gestational age; LGA, large for gestational age; LBW, low birth weight; NICU, neonatal intensive care unit.

**Table 2.**
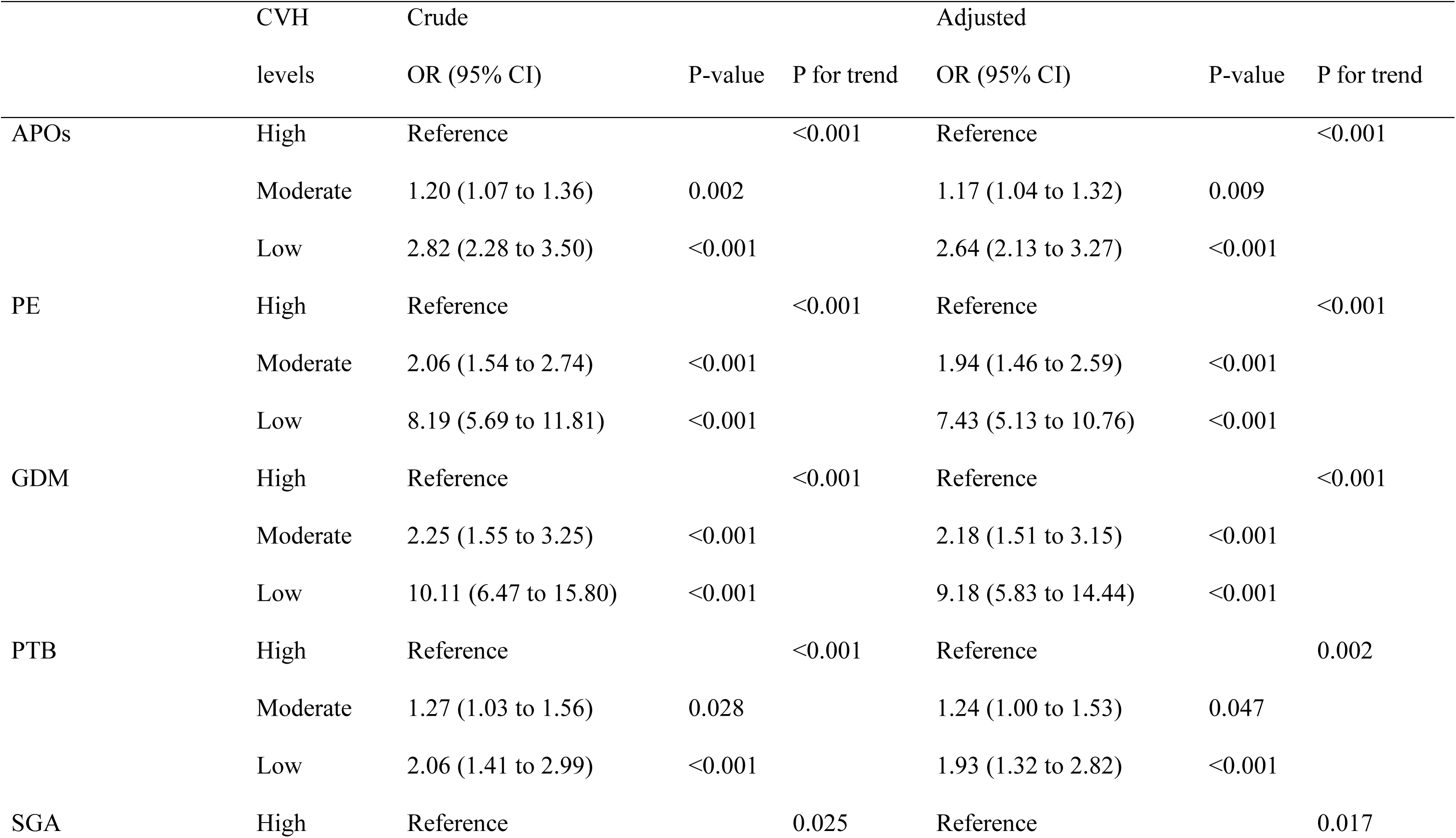
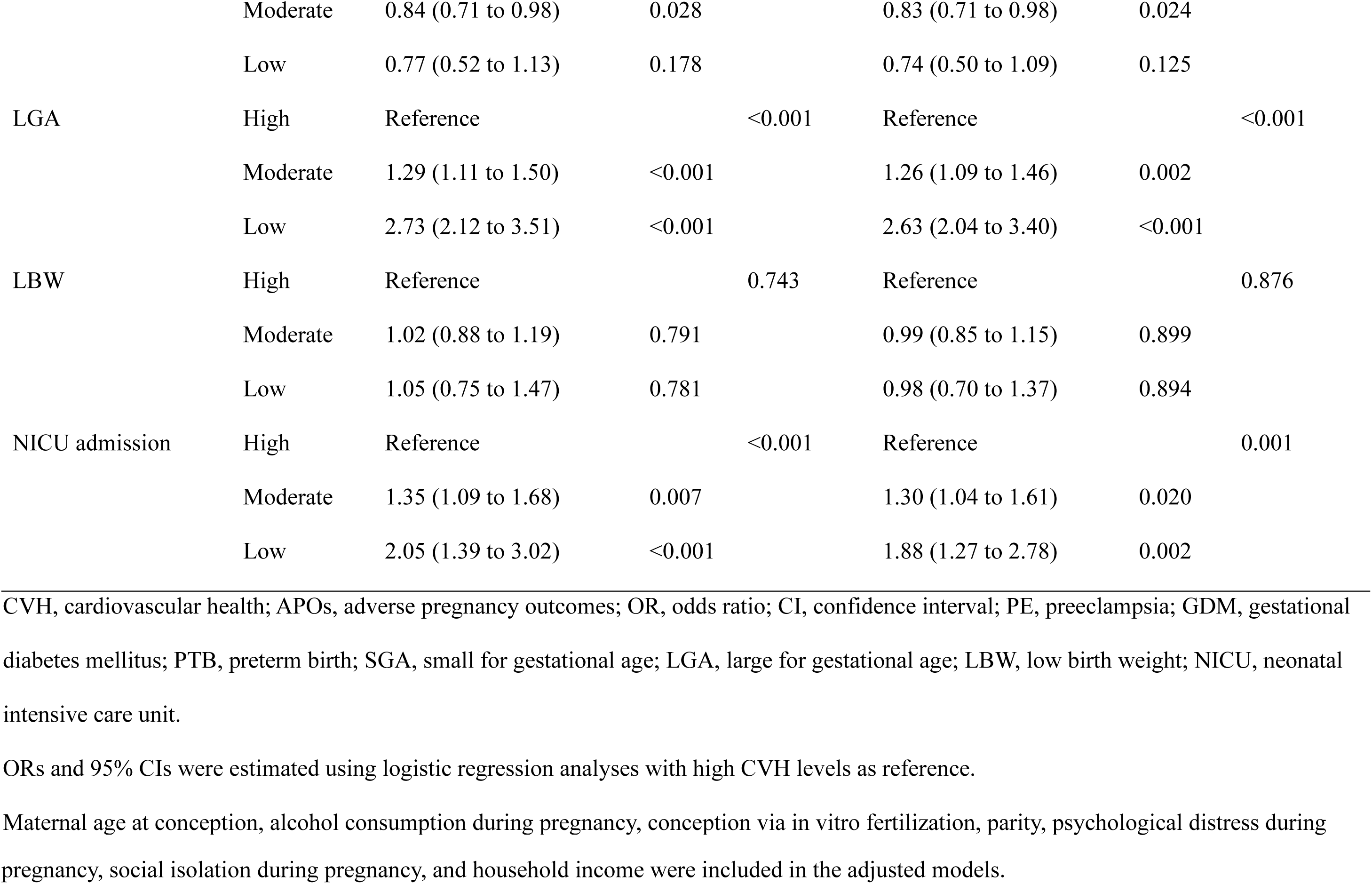
Results of logistic regression analyses showing association between overall CVH levels and study outcomes.

Subgroup analyses of psychological distress, social isolation, and income demonstrated that low overall CVH levels had a stronger association with APOs in socially isolated participants (*P* for interaction = 0.180). OR for APOs was 2.40 (95% CI: 1.86 –3.10) in those not socially isolated and 3.38 (95%CI: 2.20–5.18) in socially isolated participants (Figure 2). Among pregnant women with low CVH levels, the prevalence of APOs at delivery was higher among socially isolated pregnant women (36.4% vs. 27.4%); however, this difference was attenuated among pregnant women with high CVH levels (13.6% vs. 13.1%). Subgroup analyses of each item and subscale of social isolation were conducted, demonstrating that low overall CVH levels were more associated with APOs in participants in the low family subscale (P for interaction = 0.025). For participants with a family subscale score of ≥6, the OR (95% CI) for APOs was 2.45 (1.95–3.09) and 5.15 (2.63–10.10) in participants with a family subscale score of <6 (Figure 3). Analyses of each of the LSNS-6 items, on the family and the friend subscale, showed a significant interaction between overall CVH levels and the “Call for help” item in the family subscale.

**Figure 2.**
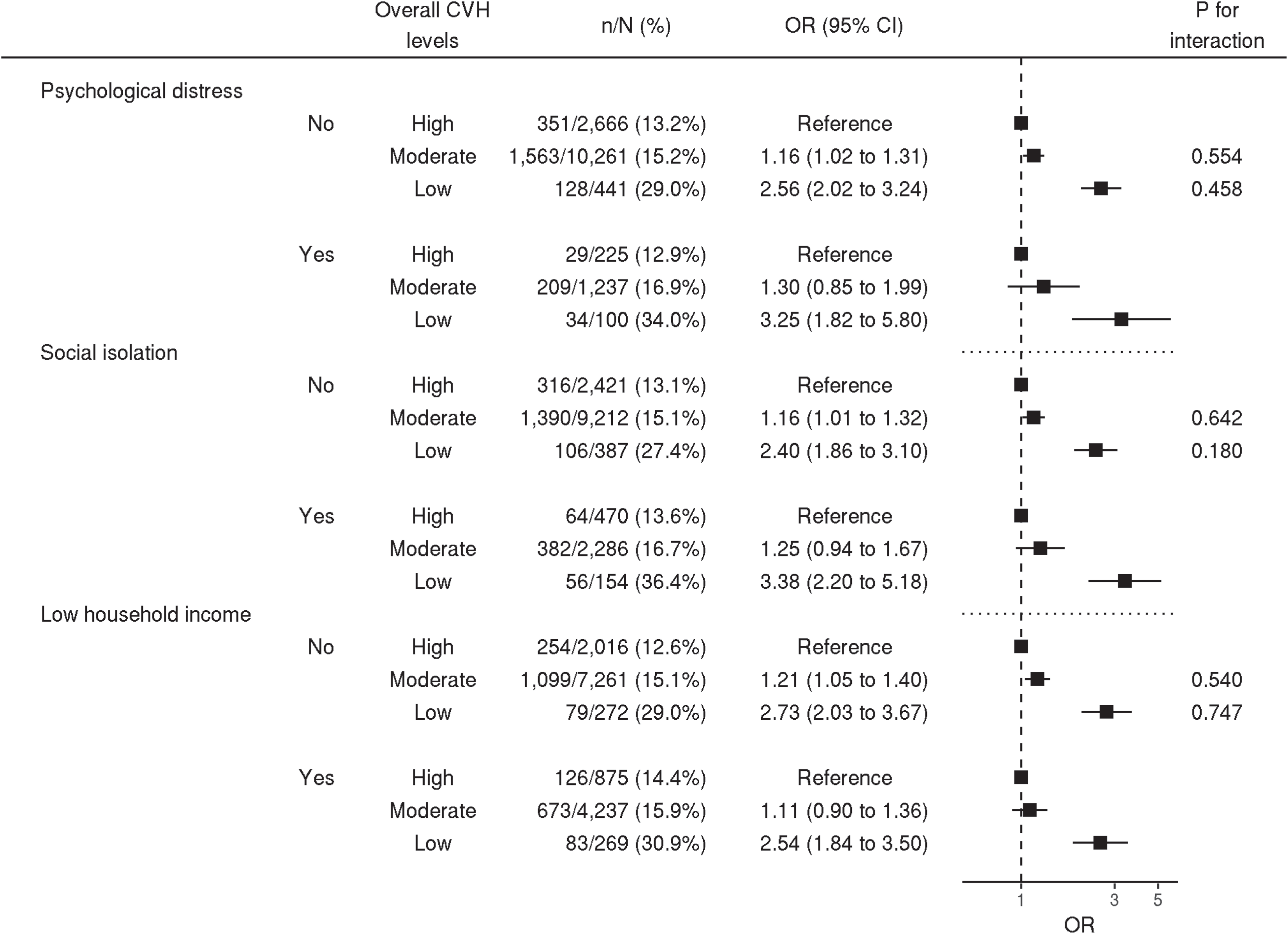
Subgroup analyses by psychological distress, social isolation, and income. ORs and 95% CIs were calculated using multiple logistic regression analyses between overall CVH levels and APOs, with adjustment for maternal age at conception, alcohol consumption during pregnancy, conception via in vitro fertilization, parity, psychological distress during pregnancy, social isolation during pregnancy, and household income. The "n/N (%)" indicates the number of cases and the total number in that stratum and its ratio. CVH: cardiovascular health; OR: odds ratio; CI confidence interval.

**Figure 3.**
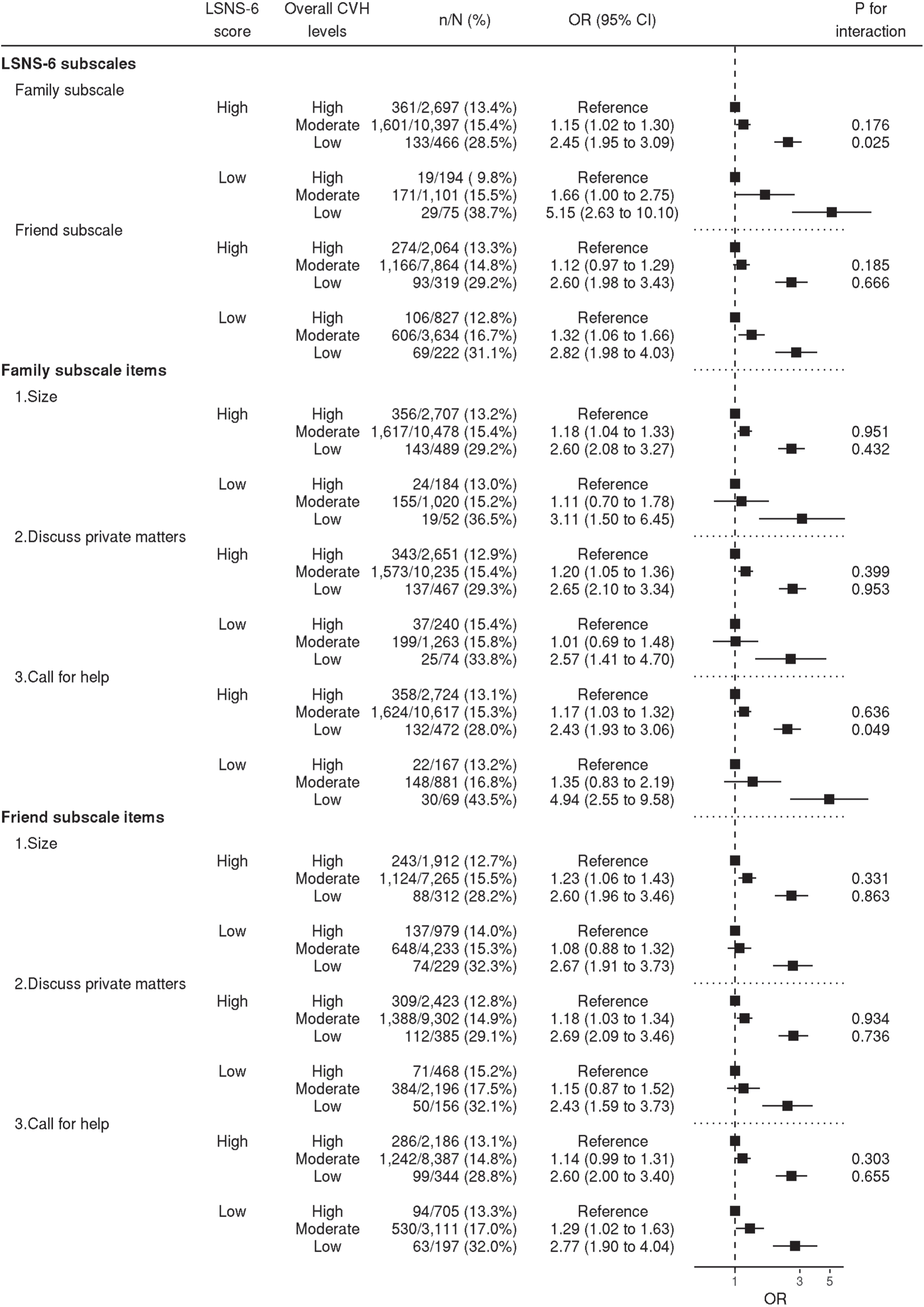
Subgroup analyses by subscales and each item in LSNS-6. The ORs and 95% CIs were calculated using multiple logistic regression analyses between overall CVH levels and APOs, with adjustment for maternal age at conception, alcohol consumption during pregnancy, conception via in vitro fertilization, parity, psychological distress during pregnancy, social isolation during pregnancy, and household income. The "n/N (%)" indicates the number of cases and the total number in that stratum and its ratio. The details of LSNS-6 were presented in Supplementary Table 2. LSNS, Lubben Social Network Scale; CVH, cardiovascular health; OR, odds ratio; CI, confidence interval.

Sensitivity analyses showed results that were largely consistent with those of the main analyses (Supplementary Figures 4-6).

### Comment

#### Principal findings

Low overall CVH levels were associated with a higher prevalence of APOs, PE, GDM, PTB, LGA, and NICU admission and a lower prevalence of SGA. The association of low overall CVH levels with APOs was stronger for socially isolated pregnant women than for pregnant women who were not socially isolated. Among the LSNS-6 items and subscales, participants with low family subscale scores and those who reported having fewer close relatives to whom they could call for help were more affected by low overall CVH levels.

#### Results in the context of what is known

Only one previous study has investigated the relationship between CVH and APOs.^19^ The previous study^19^ calculated overall CVH status using only five components and demonstrated its relationship with PE, unplanned cesarean section, LGA, sum of skinfolds, and insulin sensitivity. Our results extend these findings and further illustrate the link between overall CVH -comprising all LE8 components- and other clinical outcomes such as PTB and NICU admission. In the previous study,^19^ high overall CVH levels were positively associated with SGA, primarily through the BMI component, which was difficult to interpret and led to a clinical misinterpretation that “high CVH is a risk factor for SGA”. Our study reported similar results but also showed that a low overall CVH level is a risk factor for APOs and NICU admission. Comprehensively, our findings lend credence to the integration of CVH in antenatal care. Furthermore, the association between sleep health, a new component of LE8, and study outcomes such as GDM, SGA, and LBW suggests that LE8 may be more practical than LS7.

Our study demonstrated a more pronounced association between low overall CVH levels and APOs in socially isolated pregnant women. Negative social determinants of health, including social isolation, are well-established risk factors for CVD and mortality in non-pregnant people^37, 38^ and have been identified to influence CVH in pregnant women.^39^ However, the interaction between these factors and CVH remains elusive. Previously, a study^20^ involving middle-aged and older adults, predominantly Europeans, showed that life expectancy was comparable across socioeconomic strata among participants with optimal CVH but not among those with poor CVH, suggesting that improved CVH could mitigate health inequalities. Our findings are consistent with previous results indicating that pregnant women have a similar risk of APOs regardless of their social isolation status when their CVH improves, underscoring the importance of providing support to this population. Furthermore, our findings suggest that socially isolated pregnant women, particularly those with limited family relationships or fewer close relatives to whom they can call for help, may be more vulnerable to the impact of low CVH. Beyond the general concept of "social isolation," it may be necessary to consider the specific social conditions of pregnant women.

Notably, psychosocial distress during pregnancy did not significantly modify the effect of CVH, suggesting that psychological distress and CVH were additively related to APOs. Previous studies have reported a bidirectional relationship between CVH and psychological status, identifying maternal psychological distress as a possible risk factor for poor CVH during pregnancy.^40^ Also, improvements in CVH from early pregnancy to 6 months postpartum have been linked to reduced postpartum depressive symptoms.^41^ Psychological distress has been found to be associated with APOs.^42, 43^ Therefore, a simultaneous assessment and consideration of both aspects in clinical practice is recommended.

#### Clinical implications

Assessment of CVH as undertaken by our study has potential implications for clinical practice. Importantly, there is a need to adopt a comprehensive approach to health assessment rather than focusing solely on a specific aspect of health status. Utilizing an index of overall CVH instead of a single CVH component enables clinicians to address complex interactions among risk factors, such as smoking cessation that may lead obesity or exercise habits that may impact the duration of sleep. Particularly for socially isolated pregnant women who demonstrated a heightened vulnerability to low CVH in this study, prioritized access to primary care, lifestyle education, and access to pharmacotherapies is essential. Regardless of social isolation status, the risk of APOs was consistently low in the group with high CVH levels, suggesting that increasing CVH levels in pregnant women may reduce health disparities.

#### Research implications

Considering the correlation between CVH before and during pregnancy,^44^ incorporating LE8 in conception planning may aid in preventing future APOs. LE8 advocates improvements by lifestyle modification or the use of pharmacotherapy, empowering women to proactively enhance their CVH in preparation for pregnancy. Therefore, we hope that further study will reveal the relationship between pre-conception CVH status and APOs

#### Strength and limitations

To the best of our knowledge, this study represents the first endeavor to investigate the association between CVH assessed by LE8 and APOs. Notably, our study population was approximately five times larger than that in a prior study^19^ that assessed CVH using LS7. One strength of this study lies in the inclusion of pregnancy outcomes, providing a comprehensive assessment of the clinical importance of CVH. Also, our sensitivity analyses reinforced the robustness of the results.

Despite these strengths, our study had some limitations. First, the timing of CVH measurements varied among participants, although all assessments, including questionnaires, blood pressure measurements, and blood samples, were conducted during pregnancy. Consequently, the point at which CVH is most strongly associated with APOs remains unclear. Second, the course of pregnancy may influence behavior during pregnancy, introducing the possibility of reverse causation. Sensitivity analyses in participants with blood samples collected before or at 20 weeks showed results consistent with those of the main analyses, suggesting limited reverse causality related to glucose and lipid status. However, the effects of other conditions, such as PE diagnosis and fetal growth restriction, remain uncertain. Finally, the validity of the LE8 in the Japanese population has not yet been established. Population differences and variations in LE8 definition, such as the use of the 8-item Japanese Diet Index vs. the 16 items of Mediterranean Eating Pattern for Americans, may impact the accurate assessment of CVH by LE8.

### Conclusion

In conclusion, CVH status may be a useful index for evaluating the risk of APOs. Socially isolated pregnant women are more vulnerable to the effects of low CVH status.

## Data Availability

Individual data are available upon request after the approval of the Ethical Committee and the Materials and Information Distribution Review Committee of Tohoku Medical Megabank Organization.

## Paper presentation information

This work has not been previously presented anywhere.

## Disclaimer

None.

## Acknowledgements

The authors would like to thank all the participants who consented to participate in this study and all the staff at Tohoku Medical Megabank Organization, Tohoku University, Iwate Tohoku Medical Megabank Organization, and Iwate Medical University. A full list of the members of the Tohoku Medical Megabank Organization is available at https://www.megabank.tohoku.ac.jp/english/a230901/.

## Authors’ contributions

Conceptualization: Hisashi Ohseto, Mami Ishikuro, Geng Chen, Ippei Takahashi

Methodology: Hisashi Ohseto, Mami Ishikuro

Visualization: Hisashi Ohseto

Supervision: Shinichi Kuriyama, Taku Obara

Writing – original draft: Hisashi Ohseto, Mami Ishikuro, Geng Chen, Ippei Takahashi

Writing – review & editing: All the authors

