## Supplementary Materials for "Synergistic effects of cardiovascular health and social isolation on adverse pregnancy outcomes"



| Items | | Pages |
|---|---|---|
| Supplementary tables | | |
| | Supplementary Table 1. The CVH definition in this study | 2 |
| | Supplementary Table 2. The Lubben Social Network Scale (LSNS-6) questionnaire | 5 |
| | Supplementary Table 3. Prevalence of study outcomes by overall CVH levels | 6 |
| | Supplementary Table 4. Baseline characteristics and study outcomes of the complete cases in the TMM BirThree Cohort Study by the status of APOs | 7 |
| Supplementary figures | | |
| | Supplementary Figure 1. Flowchart showing cohort population eligible for the main analysis | 8 |
| | Supplementary Figure 2. Flowchart for sensitivity analysis in participants with blood samples collected before or at 20 weeks of gestation | 9 |
| | Supplementary Figure 3. Heatmap showing correlations between CVH components during pregnancy | 10 |
| | Supplementary Figure 4. Heatmap for sensitivity analysis showing the associations between component CVH levels and study outcomes with multiple imputation for missing values | 11 |
| | Supplementary Figure 5. Heatmap for sensitivity analysis showing the associations between component CVH levels and study outcomes in participants with blood samples collected before or at 20 weeks of gestation | 12 |
| | Supplementary Figure 6. Heatmap for sensitivity analyses showing the associations between component CVH levels and study outcomes in participants without pre-pregnancy chronic hypertension or pre-pregnancy diabetes mellitus | 13 |

**Supplementary Table 1. The CVH definition in this study**

| Components | Scoring systems |
|---|---|
| Diet | **Metrics:** The 8-item Japanese Diet Index score<br><br>**Scoring:**<br>Points Quantile<br>100 ≥95th percentile (Most adherence to the Japanese diet)<br>80 75th–94th percentile<br>50 50th–74th percentile<br>25 25th–49th percentile<br>0 1st–24th percentile (Least adherence to the Japanese diet)<br><br>**Comments:** Data for the 8-item Japanese Diet Index was collected using a food frequency questionnaire[30] based on 8 food items: rice, miso soup, seaweeds, pickles, green and yellow vegetables, fish, green tea, and red meat. For the first seven items, one point was awarded if the intake was above the median of the study population, while for red meat, one point was awarded if the intake was below the median of the study population. The total score ranged from zero to eight. Diet CVH score was calculated based on the above scoring system. |
| PA | **Metrics:** Total exercise time per week<br><br>**Scoring:**<br>Points Minutes<br>100 ≥150<br>90 120–149<br>80 90–119<br>60 60–89<br>40 30–59<br>20 1–29<br>0 0<br><br>**Comments:** During pregnancy, participants self-reported the average frequency and duration of moderate and vigorous exercise during pregnancy. One minute of moderate exercise was converted to one minute exercise and one minute of vigorous exercise was converted to two minutes of exercise. Based on this, the total exercise time per week was calculated. PA CVH score was calculated based on the above scoring system. |
| Nicotine exposure | **Metrics:** Nicotine exposure status<br><br>**Scoring:**<br>Points Status<br>100 Never smoker<br>75 Former smoker, quit ≥5 y ago<br>50 Former smoker, quit 1–<5 y ago<br>25 Former smoker, quit <1 y ago<br>0 Current smoker<br>Subtract 20 points for non-current smokers who report secondhand smoke exposure during pregnancy.<br><br>**Comments:** During pregnancy, participants self-reported current tobacco use, age of cessation, and secondhand smoke exposure during pregnancy. Inhaled nicotine-delivery system was not specifically mentioned. Nicotine exposure CVH score was calculated based on the above scoring system. |

| | |
|---|---|
| Sleep health | **Metrics:** Sleep duration without naps<br><br>**Scoring:**<br><u>Points</u> <u>Hours</u><br>100 7–<9<br>90 9–<10<br>70 6–<7<br>40 5–<6<br>20 4–<5<br>0 <4<br>Subtract 20 points from participants with sleep apnea.<br><br>**Comments:** During pregnancy, participants self-reported total daily sleep duration including naps as categorical values (<5 h, 5-<6 h, 6-<7 h, 7-<8 h, 8-<9 h, and ≥9 h) and nap duration as continuous values. The "<5 h" category was interpreted as having a continuous value of 4.5 h, and the other categories were interpreted as having values of 5.5 h, 6.5 h, 7.5 h, 8.5 h, and 9.5 h, respectively. Subsequently, sleep duration without naps was calculated by subtracting the reported nap duration from the total sleep duration including naps. History of sleep apnea was confirmed from medical record. Sleep health CVH score was calculated based on the above scoring system. |
| BMI | **Metrics:** Pre-pregnancy BMI<br><br>**Scoring:**<br><u>Points</u> <u>$kg/m^2$</u><br>100 <23.0<br>75 23.0–24.9<br>50 25.0–29.9<br>25 30.0–34.9<br>0 ≥35.0<br><br>**Comments:** Pre-pregnancy BMI was calculated from medical record. The scoring system was modified from the original definition for East Asian ancestry according to the recommendations of the American Heart Association. BMI CVH score was calculated based on the above scoring system. |
| Blood lipids | **Metrics:** Non–HDL cholesterol<br><br>**Scoring:**<br><u>Points</u> <u>mg/dL</u><br>100 <130<br>60 130–159<br>40 160–189<br>20 190–219<br>0 ≥220<br>Subtract 20 points, if drug-treated level.<br><br>**Comments:** Participants had blood samples taken during pregnancy. In the study population, all samples were taken before 32 weeks of gestation. Use of medications was self-reported during pregnancy. Blood lipids CVH score was calculated based on the above scoring system. |

| | |
|---|---|
| Blood glucose | **Metrics:** Diabetes history and HbA1c<br><br>**Scoring:**<br>Points Status<br>100 No history of diabetes and HbA1c <5.7<br>60 No diabetes and HbA1c 5.7–6.4<br>40 Diabetes with HbA1c <7.0<br>30 Diabetes with HbA1c 7.0–7.9<br>20 Diabetes with HbA1c 8.0–8.9<br>10 Diabetes with HbA1c 9.0–9.9<br>0 Diabetes with HbA1c ≥10.0<br><br>**Comments:** Participants had blood samples taken during pregnancy. In the study population, all samples were taken before 32 weeks of gestation. History of diabetes was confirmed from self-reported use of medication and medical record. Blood glucose CVH score was calculated based on the above scoring system. |
| BP | **Metrics:** Systolic and diastolic BP<br><br>**Scoring:**<br>Points mm Hg<br>100 <120/<80<br>75 120–129/<80<br>50 130–139 or 80–89<br>25 140–159 or 90–99<br>0 ≥160 or ≥100<br>Subtract 20 points, if drug-treated level.<br><br>**Comments:** Participants had their blood pressure measured during antenatal care. The first blood pressure measurement before 20 weeks of gestation was used to calculate the score. Use of medications was self-reported during pregnancy. BP CVH score was calculated based on the above scoring. |

CVH: cardiovascular health: PA: physical activity; BMI: body mass index; BP: blood pressure.

### Supplementary Table 2. The Lubben Social Network Scale questionnaire

| | |
|---|---|
| **Family subscale** | |
| 1 | **How many relatives do you see or hear from at least once a month?**<br>0 = none, 1 = one, 2 = two, 3 = three or four, 4 = five to eight, 5 = nine or more |
| 2 | **How many relatives do you feel at ease with that you can talk about private matters?**<br>0 = none, 1 = one, 2 = two, 3 = three or four, 4 = five to eight, 5 = nine or more |
| 3 | **How many relatives do you feel close to such that you could call on them for help?**<br>0 = none, 1 = one, 2 = two, 3 = three or four, 4 = five to eight, 5 = nine or more |
| Friend subscale | |
| 1 | **How many friends do you see or hear from at least once a month?**<br>0 = none, 1 = one, 2 = two, 3 = three or four, 4 = five to eight, 5 = nine or more |
| 2 | **How many friends do you feel at ease with that you can talk about private matters?**<br>0 = none, 1 = one, 2 = two, 3 = three or four, 4 = five to eight, 5 = nine or more |
| 3 | **How many friends do you feel close to such that you could call on them for help?**<br>0 = none, 1 = one, 2 = two, 3 = three or four, 4 = five to eight, 5 = nine or more |

The Lubben Social Network Scale score was calculated by summing all six items. Total score ranged from zero to 30. Scores of <12 were considered socially isolated.

The family and friend subscales were calculated by summing each of the three items and the total ranged from zero to 15. Scores below six were considered socially isolated.

### Supplementary Table 3. Prevalence of study outcomes by overall CVH levels

| | High overall CVH | Moderate overall CVH | Low overall CVH | P-value |
|---|---|---|---|---|
| | n = 2,891 | n = 11,498 | n = 541 | |
| APOs, % | 380 (13.1) | 1,772 (15.4) | 162 (29.9) | <0.001 |
| PE, % | 54 (1.9) | 433 (3.8) | 73 (13.5) | <0.001 |
| GDM, % | 32 (1.1) | 282 (2.5) | 55 (10.2) | <0.001 |
| PTB, % | 108 (3.7) | 539 (4.7) | 40 (7.4) | 0.001 |
| SGA, % | 213 (7.4) | 718 (6.2) | 31 (5.7) | 0.071 |
| LGA, % | 230 (8.0) | 1,155 (10.0) | 103 (19.1) | <0.001 |
| LBW, % | 225 (7.8) | 912 (7.9) | 44 (8.1) | 0.947 |
| NICU admission, % | 100 (3.5) | 531 (4.6) | 37 (6.8) | 0.001 |

CVH, cardiovascular health; APOs, adverse pregnancy outcomes; PE, preeclampsia; GDM, gestational diabetes mellitus; PTB, preterm birth; SGA, small for gestational age; LGA, large for gestational age; LBW, low birth weight; NICU, neonatal intensive care unit.
Data are presented as n (%). P-values were calculated using the chi-square test.

**Supplementary Table 4. Baseline characteristics and study outcomes of the complete cases in the TMM BirThree Cohort Study by the status of APOs**

| | With APOs n = 2,314 | Without APOs n = 12,616 | P-value |
|---|---|---|---|
| Advanced maternal age at conception, % | 710 (30.7) | 3,214 (25.5) | <0.001 |
| Low income, % | 882 (38.1) | 4,499 (35.7) | 0.025 |
| Alcohol consumption during pregnancy, % | 461 (19.9) | 2,513 (19.9) | >0.999 |
| Psychological distress, % | 272 (11.8) | 1,290 (10.2) | 0.030 |
| Social isolation, % | 502 (21.7) | 2,408 (19.1) | 0.004 |
| Conception by IVF, % | 147 (6.4) | 607 (4.8) | 0.002 |
| Primipara, % | 904 (39.1) | 4,982 (39.5) | 0.719 |
| GA at blood sampling, day | 137.2 ± 34.4 | 136.1 ± 33.3 | 0.150 |
| GA at BP measurement, day | 79.3 ± 15.8 | 79.4 ± 15.2 | 0.778 |
| **CVH scores** | | | |
| Overall score | 68.1 ± 11.9 | 70.6 ± 10.6 | <0.001 |
| Diet score | 48.8 ± 33.1 | 48.3 ± 33.2 | 0.529 |
| PA score | 23.3 ± 39.7 | 24.7 ± 40.5 | 0.127 |
| Nicotine exposure score | 69.0 ± 35.0 | 72.1 ± 33.1 | <0.001 |
| Sleep health score | 71.4 ± 27.7 | 73.0 ± 26.5 | 0.008 |
| BMI score | 82.3 ± 30.2 | 87.1 ± 25.2 | <0.001 |
| Blood lipids score | 68.3 ± 29.6 | 69.0 ± 29.5 | 0.243 |
| Blood glucose score | 97.8 ± 10.5 | 99.4 ± 5.2 | <0.001 |
| BP score | 83.6 ± 23.6 | 91.1 ± 17.1 | <0.001 |
| **Outcomes** | | | |
| PE, % | 560 (24.2) | 0 (0.0) | <0.001 |
| GDM, % | 369 (15.9) | 0 (0.0) | <0.001 |
| PTB, % | 687 (29.7) | 0 (0.0) | <0.001 |
| SGA, % | 962 (41.6) | 0 (0.0) | <0.001 |
| LGA, % | 176 (7.6) | 1,312 (10.4) | <0.001 |
| LBW, % | 879 (38.0) | 302 (2.4) | <0.001 |
| NICU admission, % | 376 (16.2) | 292 (2.3) | <0.001 |

APOs, adverse pregnancy outcome; IVF, in vitro fertilization; GA, gestational age; BP, blood pressure; CVH, cardiovascular health; PA, physical activity; BMI, body mass index; PE, preeclampsia; GDM, gestational diabetes mellitus; PTB, preterm birth; SGA, small for gestational age; LGA, large for gestational age; LBW, low birth weight; NICU, neonatal intensive care unit.
Data are shown as mean ± standard deviation for continuous variables and n (%) for categorical variables.
P-values were calculated using Student's t-test for continuous variables and the chi-square test for categorical variables.

### Online-only figures

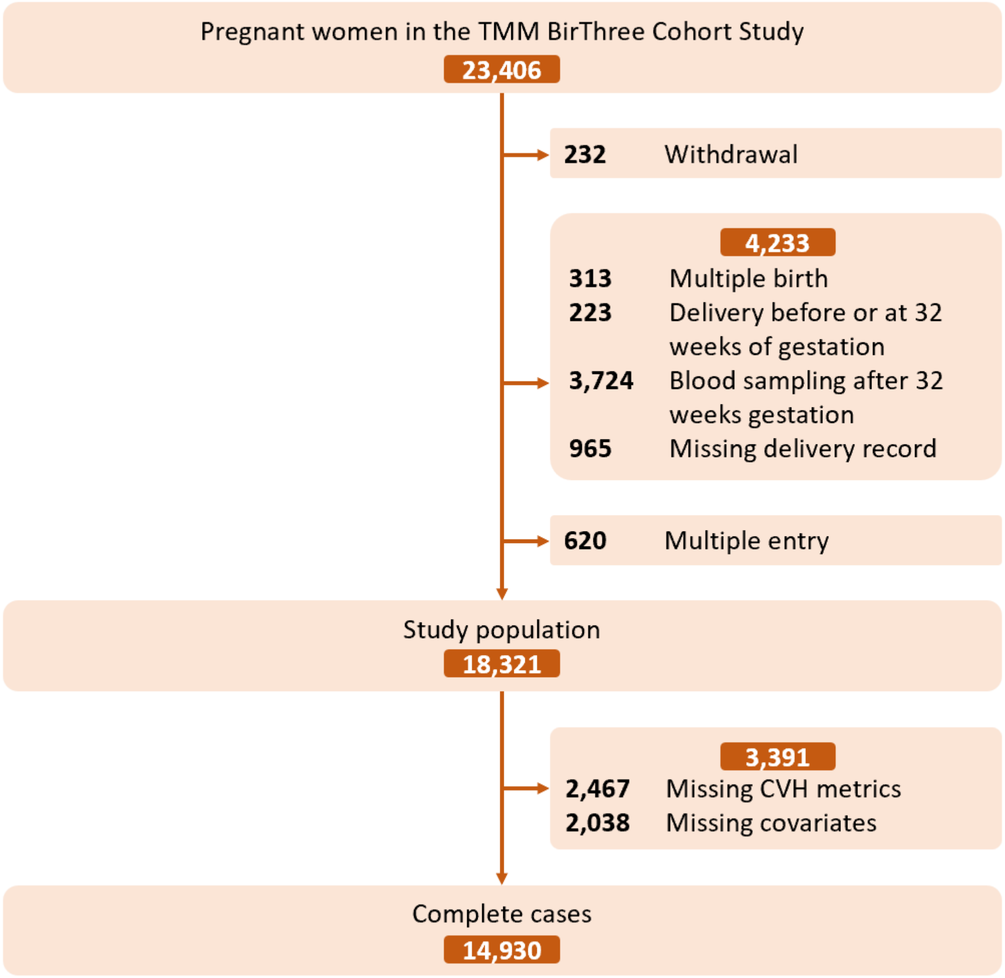


**Supplementary Figure 1. Flowchart showing cohort population eligible for the main analysis**

A total of 18,321 participants fulfilled the eligibility criteria, and the main analysis involved 14,930 participants with complete data.
TMM BirThree Cohort Study, Tohoku Medical Megabank Project Birth and Three-Generation Cohort Study; CVH, cardiovascular health.

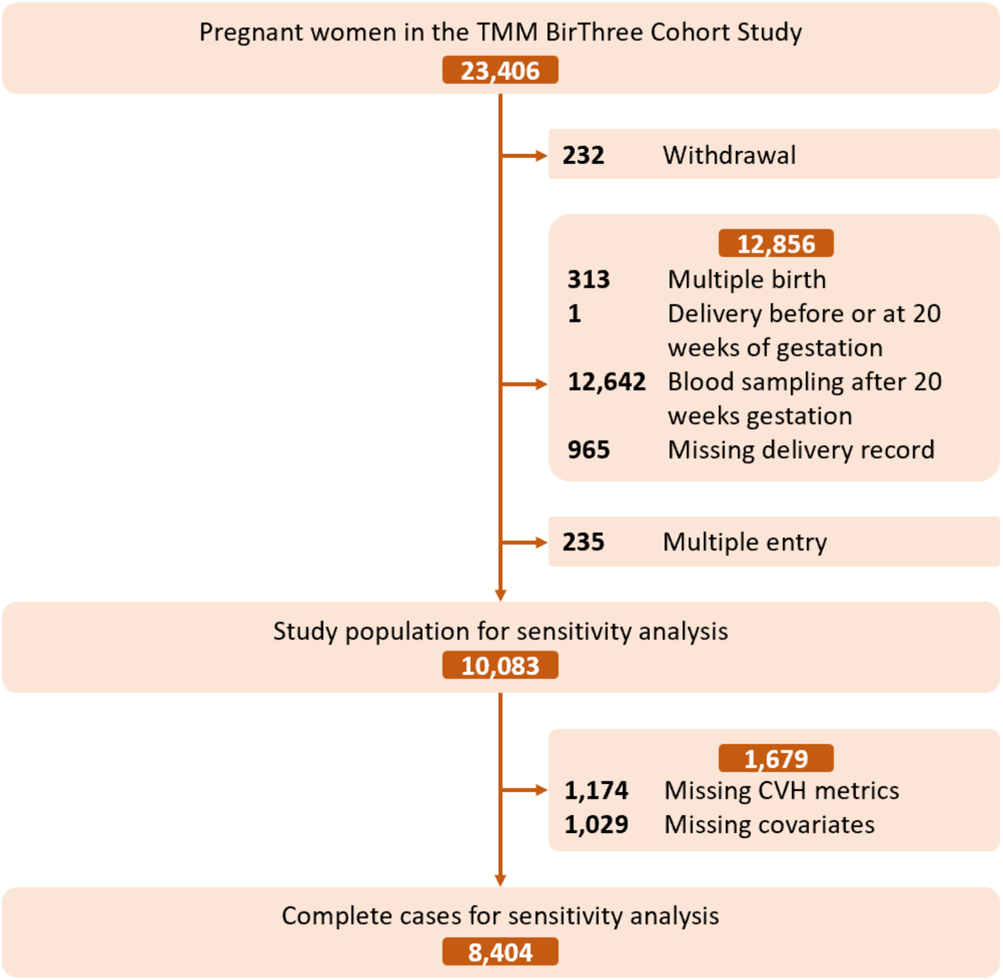


**Supplementary Figure 2. Flowchart for sensitivity analysis in participants with blood samples collected before or at 20 weeks of gestation**

Sensitivity analysis was conducted for 8,404 participants with blood samples collected before or at 20 weeks.

TMM BirThree Cohort Study, Tohoku Medical Megabank Project Birth and Three-Generation Cohort Study; CVH. cardiovascular health.

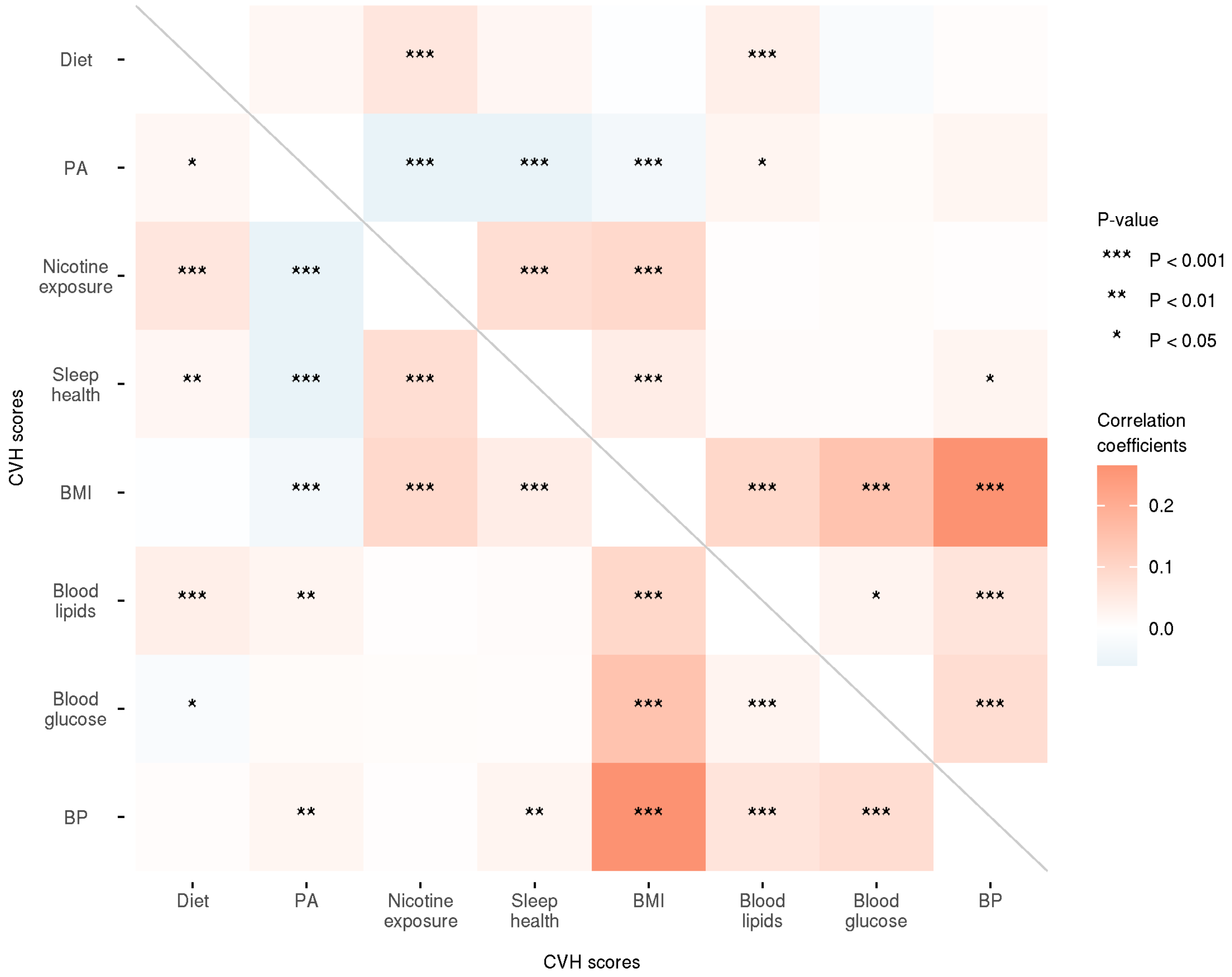


**Supplementary Figure 3. Heatmap showing correlations between CVH components during pregnancy**

The heatmap shows Pearson's correlation coefficients between CVH scores for individual components. Red, blue, and white indicate positive, negative, and no association, respectively. Darker colors indicate stronger associations. The asterisk indicates the p-value. CVH, cardiovascular health; PA, physical activity; BMI, body mass index; BP, blood pressure.

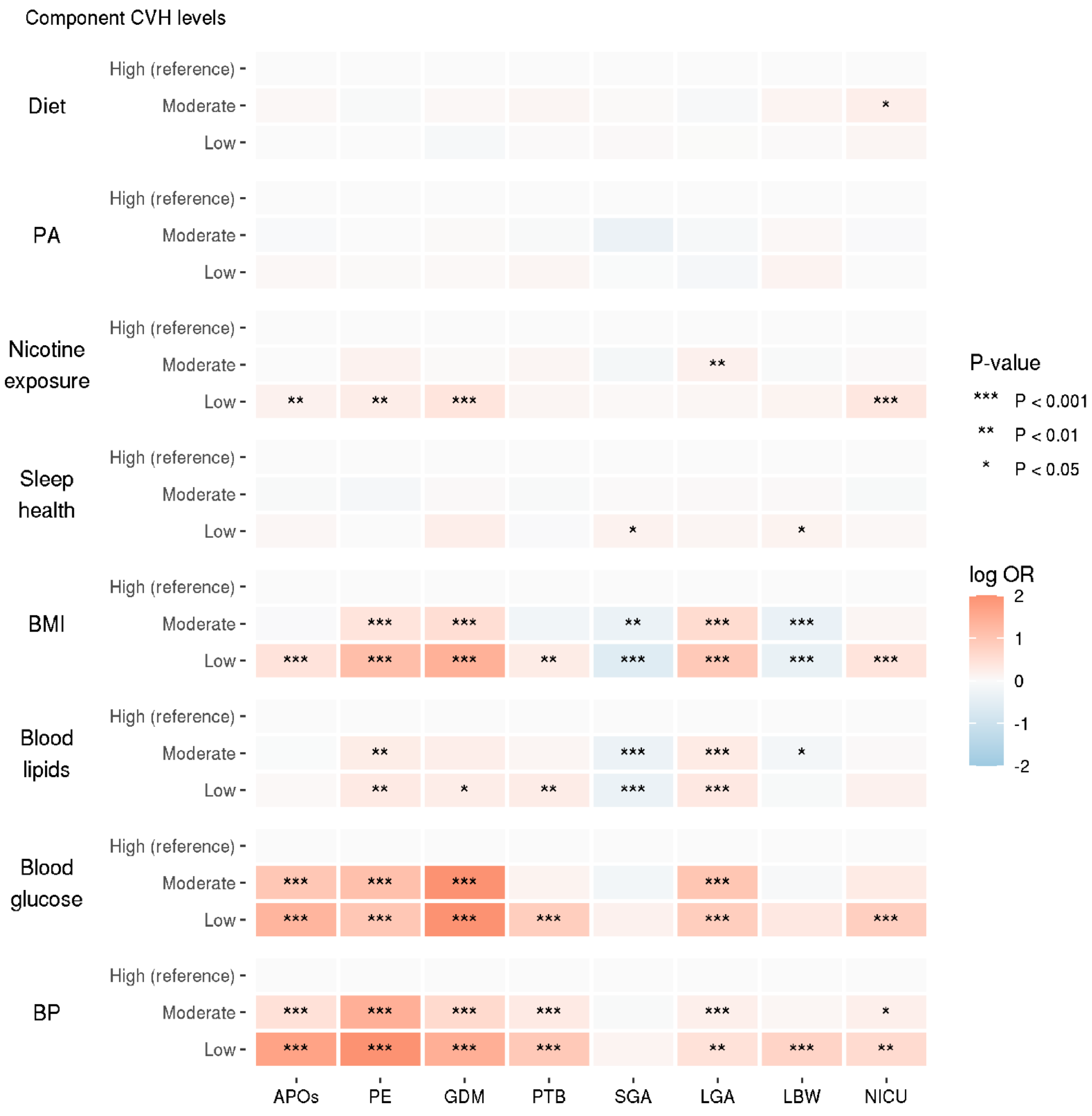


**Supplementary Figure 4. Heatmap for sensitivity analysis showing the associations between component CVH levels and study outcomes with multiple imputation for missing values**

The heatmap represents log transformed odds ratios (ORs) from multiple logistic regression of sensitivity analysis conducted with multiple imputations for missing values. Adjustments were made for maternal age at conception, alcohol consumption during pregnancy, conception via in vitro fertilization, parity, psychological distress during pregnancy, social isolation during pregnancy, and household income. High CVH levels were reference s. Red, blue, and grey indicate positive, negative, and no association, respectively. Darker colors indicate stronger associations. The asterisk indicates the p-value. For clarity, extreme Log OR values are presented as 2 or -2 when their absolute values are 2 or more.

CVH, cardiovascular health; OR, odds ratio; PA, physical activity; BMI, body mass index; BP, blood pressure; APOs, adverse pregnancy outcomes; PE, preeclampsia; GDM, gestational diabetes mellitus; PTB, preterm birth; SGA, small for gestational age; LGA, large for gestational age; LBW, low birth weight; NICU, neonatal intensive care unit.

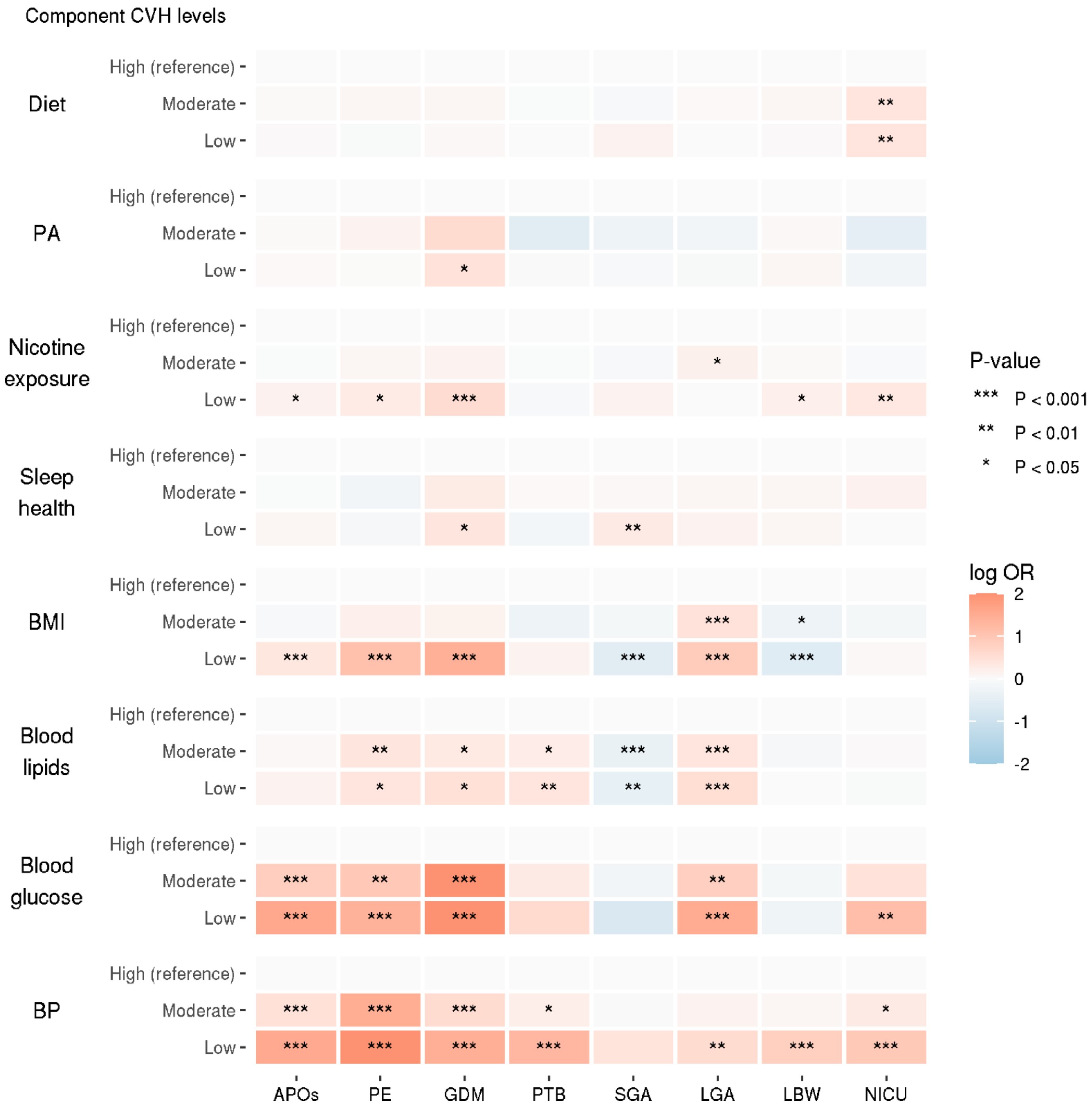


**Supplementary Figure 5. Heatmap for sensitivity analysis showing the associations between component CVH levels and study outcomes in participants with blood samples collected before or at 20 weeks of gestation**

The heatmap represents log transformed odds ratios (ORs) from multiple logistic regression of sensitivity analysis conducted in participants with blood samples collected before or at 20 weeks of gestation. Adjustments were made for maternal age at conception, alcohol consumption during pregnancy, conception via in vitro fertilization, parity, psychological distress during pregnancy, social isolation during pregnancy, and household income. High CVH levels were references. Red, blue, and grey indicate positive, negative, and no association, respectively. Darker colors indicate stronger associations. The asterisk indicates the p-value. For clarity, extreme Log OR values are presented as 2 or -2 when their absolute values are 2 or more.

CVH, cardiovascular health; OR, odds ratio; PA, physical activity; BMI, body mass index; BP, blood pressure; APOs, adverse pregnancy outcomes; PE, preeclampsia; GDM, gestational diabetes mellitus; PTB, preterm birth; SGA, small for gestational age; LGA, large for gestational age; LBW, low birth weight; NICU, neonatal intensive care unit.

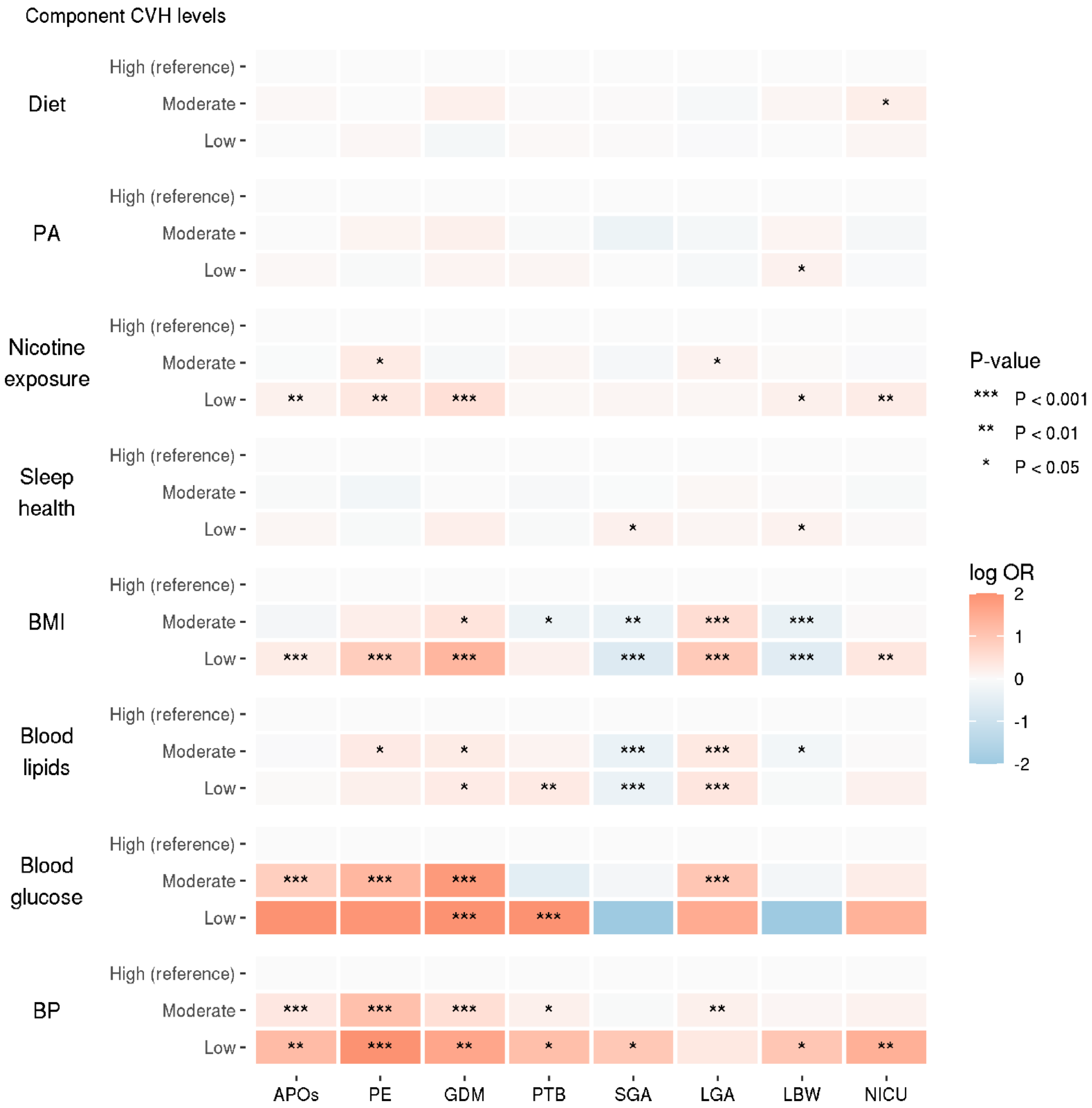


**Supplementary Figure 6. Heatmap for sensitivity analyses showing the associations between component CVH levels and study outcomes in participants without pre-pregnancy chronic hypertension or pre-pregnancy diabetes mellitus**

The heat map represents log transformed odds ratios (ORs) from multiple logistic regression of sensitivity analysis in participants without pre-pregnancy chronic hypertension or pre-pregnancy diabetes mellitus. Adjustments were made for maternal age at conception, alcohol consumption during pregnancy, parity, conception via in vitro fertilization, psychological distress during pregnancy, social isolation during pregnancy, and household income. High CVH levels were used as reference values. Red, blue, and grey indicate positive, negative, and no association, respectively. Darker colors indicate stronger associations. The asterisk indicates the p-value. For clarity, extreme Log OR values are presented as 2 or -2 when their absolute values are 2 or more.

CVH, cardiovascular health; OR, odds ratio; PA, physical activity; BMI, body mass index; BP, blood pressure; APOs, adverse pregnancy outcomes; PE, preeclampsia; GDM, gestational diabetes mellitus; PTB, preterm birth; SGA, small for gestational age; LGA, large for gestational age; LBW, low birth weight; NICU, neonatal intensive care unit.
